# Brain predictors of fatigue in Rheumatoid Arthritis: a machine learning study

**DOI:** 10.1101/2021.10.15.21265049

**Authors:** María Goñi, Neil Basu, Alison D. Murray, Gordon D. Waiter

## Abstract

**Background:** Fatigue is a common and burdensome symptom in Rheumatoid Arthritis (RA), yet is poorly understood. Currently, clinicians rely solely on fatigue questionnaires, which are inherently subjective measures. For the effective development of future therapies and stratification, it is of vital importance to identify biomarkers of fatigue. In this study, we identify brain differences between RA patients who improved and did not improve their levels of fatigue, and we compared the performance of different classifiers to distinguish between these samples at baseline.

**Methods:** Fifty-four fatigued RA patients underwent a magnetic resonance (MR) scan at baseline and 6 months later. At 6 months we identified those whose fatigue levels improved and those for whom it did not. More than 900 brain features across three data sets were assessed as potential predictors of fatigue improvement. These data sets included clinical, structural MRI (sMRI) and diffusion tensor imaging (DTI) data. A genetic algorithm was used for feature selection. Three classifiers were employed in the discrimination of improvers and non-improvers of fatigue: a Least Square Linear Discriminant (LSLD), a linear Support Vector Machine (SVM) and a SVM with Radial Basis Function kernel. Results: The highest accuracy (67.9%) was achieved with the sMRI set, followed by the DTI set (63.8%), whereas classification performance using clinical features was at the chance level. The mean curvature of the left superior temporal sulcus was most strongly selected during the feature selection step, followed by the surface are of the right frontal pole and the surface area of the left banks of the superior temporal sulcus. Conclusions: The results presented in this study evidence a superiority of brain metrics over clinical metrics in predicting fatigue changes. Further exploration of these methods may enable clinicians to triage patients towards the most appropriate fatigue alleviating therapies.

## Introduction

Fatigue is a highly complex symptom, which is challenging to measure and study. There are around 690,000 RA patients in UK, 80% of whom report suffering from relevant fatigue(1). This symptom is described as a principal burden of the disease (2), with an impact over the quality of life even greater than pain(3). Consequences of fatigue include an impact upon daily activities, general health, mental health, work and relationships among others (3).

Therapeutic options do exist. In particular, exercise, cognitive behavioural therapy (CBT) and education are recommended; moreover, we have recently evidenced a fatigue alleviating effect of anti-TNF pharmacological therapy(4). However, in practice, these interventions only provide clinically meaningful gains in subgroups of patients (5,6). Unfortunately, clinical characteristics inadequately distinguish these subgroups, knowledge which would otherwise inform therapeutic decision making. Consequently, patients and clinicians are frustrated by the considerable challenge of selecting the correct therapy for the correct patient.

Given that central symptoms appear prominently associated with fatigue (7), neuroimaging may be a valuable tool to study fatigue and potentially identify biomarkers which may guide clinician’s therapeutic decision making. In spite of this possibility, few studies have employed these techniques to investigate fatigue in the field of chronic inflammatory diseases (8). Yet, these studies are restricted to traditional univariate analysis (for instance t-test, chi-square or Wilcoxon test) to test for differences between groups. Therefore, these findings are group-based and hence of limited clinical application. In addition, no studies have employed neuroimaging to predict fatigue outcome. This information would be a vital asset in stratification of therapy, currently undoable as clinicians rely only on clinical measures.

Machine learning (ML) methods integrated with neuroimaging are gaining importance to support clinical diagnosis. As opposed to group level comparison analysis, machine learning methods enable identification of fine-grained patterns, which are key for single subject diagnosis (9). This work is the first study to apply machine learning techniques in the search for brain indicators which may predict longitudinal changes in fatigue and so aid patient stratification in patients with RA.

## Methods

### Participants

The data used in this work was acquired in Aberdeen Biomedical Imaging Centre. The main eligibility criteria for enrolment include: a) patients diagnosed with RA, according to the 2010 American College of Rheumatology/European League Against Rheumatism (ACR/EULAR) criteria (10) and b) patients suffering from clinically relevant fatigue, defined as a score above 3 on the Chalder Fatigue Scale (CFS)(11) and reported fatigue for more than 3 months. Those patients with contraindications to MRI scanning (e.g. pacemakers, artificial eyes, artificial joints), claustrophobia, alternative medical justifications for their fatigue (e.g. beta-blocker treatment, a recent history of cancer) and left-handed patients were excluded. Finally, 60 patients met eligibility criteria and thus, underwent a full clinical assessment, before going through the MR scan. The whole procedure was repeated after 6 months, with no fatigue specific intervention in the meanwhile, in 54 out of the 60 initial patients who returned for a second visit. In the second visit, patients were stratified based on Chalder Fatigue Scale score variation (ΔCFS) into improvers (ΔCFS ≥ 2; n = 22; mean age 54.9 ± 13.5; 15 female) and non-improvers of fatigue (ΔCFS < 2; n = 32; mean age 55 ± 10.0; 26 female). Standard clinical characteristics are provided in table 1. Blood and questionnaire-related measures are summarized in **Table S1**.

**Table 1.**
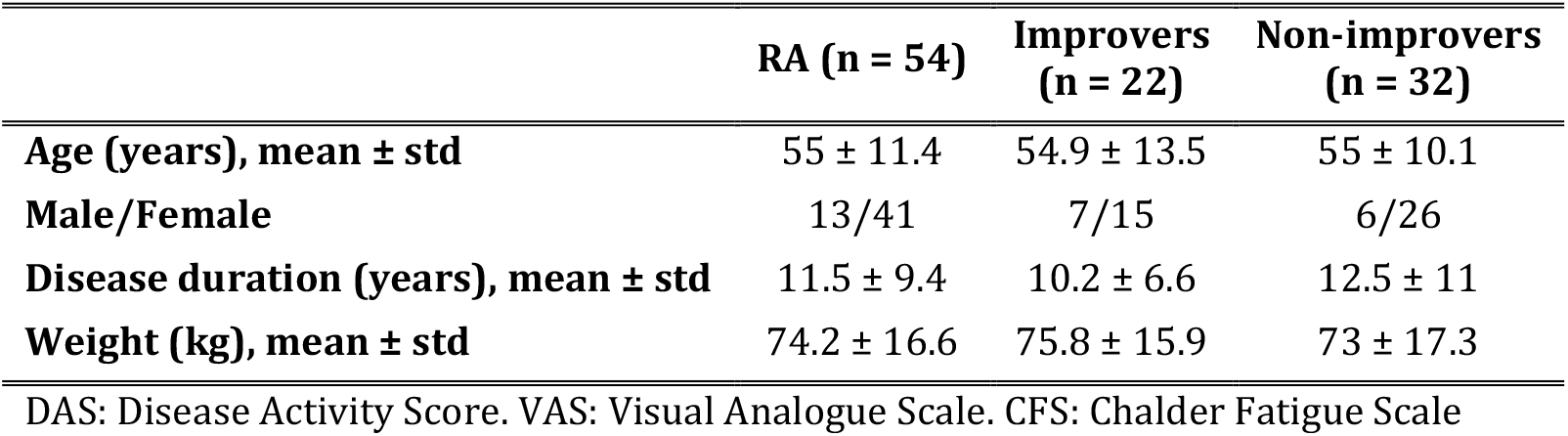
Clinical characteristics for the total number of subjects (n=54) and improvers (n=22) and non-improvers (n=32) of fatigue subgroups.

|  | RA (n = 54) | Improvers (n = 22) | Non-improvers (n = 32) |
| --- | --- | --- | --- |
| **Age (years), mean ± std** | 55 ± 11.4 | 54.9 ± 13.5 | 55 ± 10.1 |
| **Male/Female** | 13/41 | 7/15 | 6/26 |
| **Disease duration (years), mean ± std** | 11.5 ± 9.4 | 10.2 ± 6.6 | 12.5 ± 11 |
| **Weight (kg), mean ± std** | 74.2 ± 16.6 | 75.8 ± 15.9 | 73 ± 17.3 |

DAS: Disease Activity Score. VAS: Visual Analogue Scale. CFS: Chalder Fatigue Scale

### Data acquisition

Patients were asked to lie supine in a 3 Tesla Philips Achieva X-series MRI scanner for a brain scan. Structural MRI data was acquired by a T1-weighted fast-field echo 3D structural scan with the next parameters: repetition time (TR) = 8.2 ms, echo time (TE) = 3.8 ms, inversion time (TI) = 1018 ms, flip angle (FA) = 8°, field of view (FOV) = 240 mm, matrix size = 240 × 240 with 160 slices, voxel size = 0.94 × 0.94 × 1 mm^3^.

Diffusion tensor images (DTI) were acquired along 16 gradients directions (b = 800 s/mm^2^, number of excitations = 2) along with an unweighted (b = 0) image with an overall of 17 volumes. DTI images were recorded as a series of 66 axial slices, using a single-shot spin echo planar imaging (EPI) sequence with the next parameters: TR = 7151 ms, TE = 55 ms, FA = 90°, FOV = 224 × 224 mm^2^, slice thickness = 2.0 mm with no gap, voxel size = 2 × 2 × 2 mm^3^, matrix size = 224 × 224 × 132.

### Preprocessing

T1-weighted images were pre-processed using FreeSurfer. Some of the processing steps include: 1) motion correction and averaging, 2) registration to the Talairach coordinates, 3) intensity normalization, 4) skull stripping using a hybrid watershed/surface deformation algorithm to remove brain tissue from non-brain tissue such as skull, eyeballs and skin, 5) automatic volume labelling to assign each voxel to one of 57 regions of interest, 6) intensity normalization using just the brain volume as the input, 7) white matter segmentation to separate white matter from anything else, 8) fill and cut subcortical mass to fill in any holes, remove any islands and brain stem and separate hemispheres, 9) surface between grey and white matter, and the pial surface are generated and refined.

DTI data were pre-processed using a Tract-Based Automatic Analysis (TBAA). First, diffusion tensor imaging (DTI) data were registered to create a single study specific template (SST). This SST tackles the bias from group differences. Then, the SST was registered to the NTU-DSI-122 template, which gathers a total of 76 fiber tracts. After that, sampling coordinates were transformed by reversing the process, from the NTU-DSI-122 template to each individual DTI dataset. Finally, the generalized FA (GFA), a measure of the microstructural differences of the 76 fiber tracts, is sampled according to the transformed coordinates in native space.

### Feature extraction

As a result of the pre-processing step, 52 subcortical measures and 68 cortical parcellations (34 per hemisphere) were extracted (**Table S2**). For each of the subcortical segmentations, volume in mm^3^ were calculated. In addition, eight statistic measures were calculated for each of the cortical parcellations, including: 1) total surface area (mm^2^), 2) total gray matter volume (mm^3^), 3) average cortical thickness (mm), 4) standard deviation of cortical thickness (mm), 5) integrated rectified mean curvature (mm^-1^), 6) integrated rectified gaussian curvature (mm^-2^), 7) folding index (unitless) and 8) intrinsic curvature index (unitless). An overall of 596 structural features (52 subcortical and 544 cortical) were calculated.

Each of 76 fiber tracts or diffusion tensors extracted in the pre-processing step can be described by 3 eigenvalues, expressing magnitude, degree and orientation of diffusion anisotropy. For each diffusion tensor, 4 parameters were calculated based on the eigenvalues: fractional anisotropy, mean diffusivity, longitudinal diffusivity and radial diffusivity. As a result of the 4 aforementioned measures for each of the 76 diffusion tensors (**Table S3**), 304 diffusion features were gathered.

### Feature selection

As the dimensionality of the data grows, data analysis and prediction algorithms become increasingly complex or even incorrect (12). The final aim of feature selection is to select the best subset of features in a certain prediction problem, reducing dimensionality, improving the learning performance and decreasing the computational cost. A genetic algorithm (GA)(13) was employed to identify a subset of features able to distinguish between improvers and non-improvers of fatigue. GAs are inspired by Charles Darwin’s theory of natural evolution, where the fittest individuals from a population survive, having more chances to reproduce and then, transmit their genetic material to their offspring. This process is repeated until reaching a generation with the fittest individuals.

This process can be extrapolated to a feature selection problem. Here, an individual of the population represents a possible solution to the prediction problem (optimal subset of features). Each individual is characterized by a set of genes (features). Genes are gathered in a string known as chromosome (possible solution). A GA conceives new generations of the population, where individuals offer a better solution to the problem each time. The algorithm performs the following steps (Figure 1, B):

**Figure 1.**
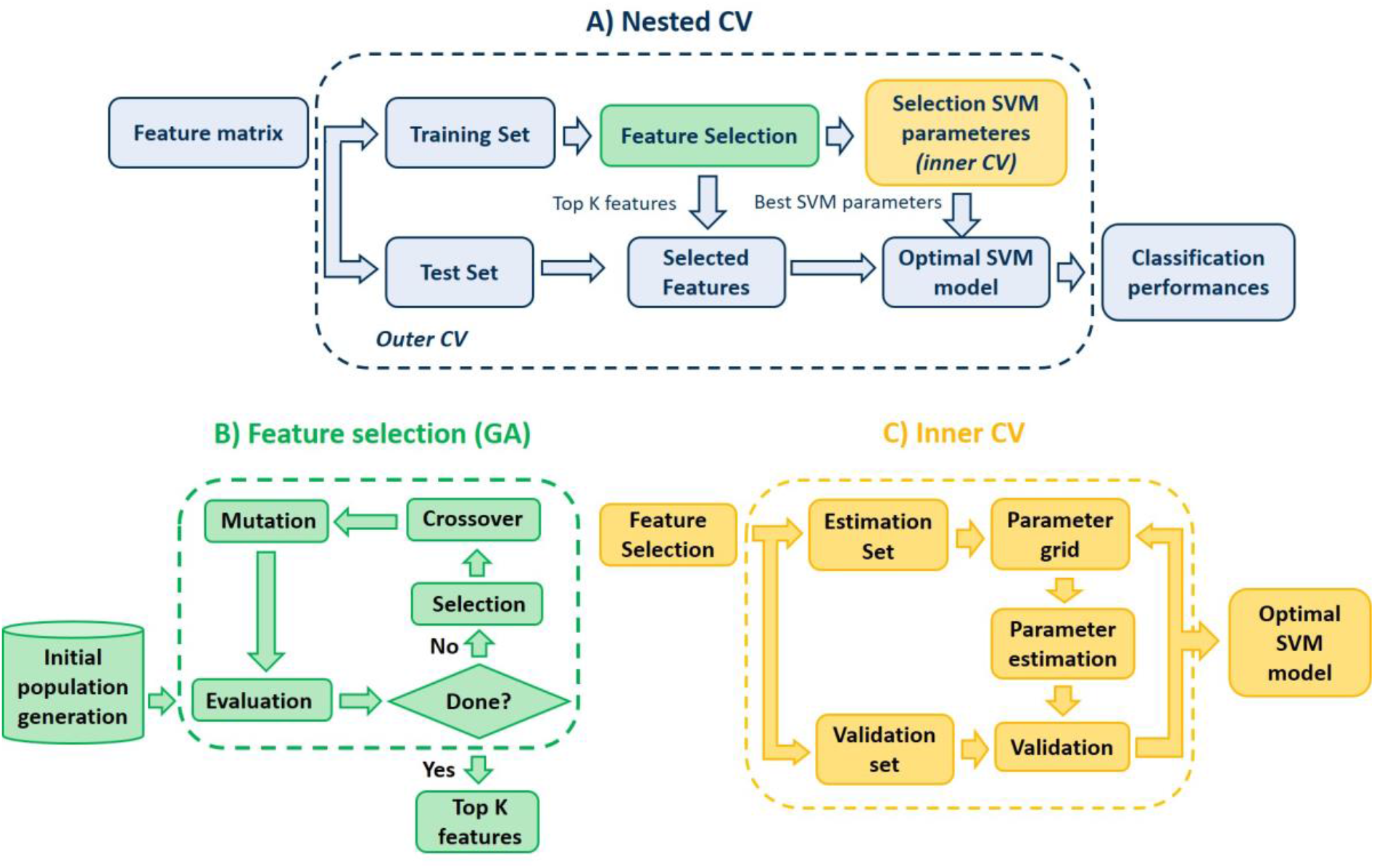
(A) Nested CV procedure for model development and evaluation. The outer loop consists of a 5-fold CV, used to evaluate the final model on the held out test set. This CV procedure is repeated 100 times. (B) The feature selection step consists of a genetic algorithm to select the optimal features in the classification problem. (C) The inner loop, consists of a 5-fold CV, employed to tune the parameters of the SVM.

1. Initial population. The process starts selecting a set of chromosomes that defines the initial population. A population size of 10 times the number of dimensions was stablished(14).
2. Fitness function. This function is defined to evaluate the probability of the individual to be chosen for reproduction. Fitter chromosomes will have a higher fitness value. A Least Squared Linear Discriminant (LSLD) was employed in this work. Then, in each iteration, the GA selects the feature combination which minimizes the mean squared error of the LSLD over the training data.
3. Selection operator. During the selection step, *m* individuals (parents) are chosen for reproduction based on their fitness scores. There are several selection methods (15). Here, a ranking-based approach is employed, where chromosomes are arranged according to their fitness values. A subset of 10% of the population is selected.
4. Crossover operator. The aim is to transfer the genetic material from generation to generation (16). The new offspring inherits genes from both “parents”. A random crossover approach is the selected method in this work (17).
5. Mutation operator. This operator allows a wide search space by changing randomly a certain number of bits. In this work, 1% of the genes were randomly mutated.
6. Termination criterion: The whole process is repeated until a certain termination criterion is reached. Here, the process terminates after 100 generations, where the optimal combination of features is returned.

### Classification

Classification was performed using three different classifiers to distinguish between improvers and non-improvers of fatigue: a least square linear discriminant (LSLD), a soft-margin SVM and a Gaussian SVM.

#### Least Square Linear Discriminant

The aim of a Linear Discriminant Analysis (LDA) is to combine *L* features of a dataset of *N* instances, to distinguish effectively between *C* classes. The decision function can be expressed as follows:

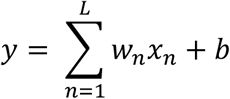

Where *x* is the input vector, *b* is a bias term and *w* is a feature weight vector, which represents the contribution of each feature to the prediction. The Least Squares Linear Discriminant (LSLD) is optimized by minimizing the mean square error (18).

#### Support Vector Machines

A support vector machine (SVM) classifier seeks to find a hyperplane with the maximum distance between the closest points to the hyperplane itself (19). Then, this hyperplane is just defined by the training instances, which fall just in the frontier of the margin, known as support vectors. Originally, SVM was a linear algorithm, but modifications to deal with non-linearly separable classes have been introduced. These modifications are the ones employed in this work and include the soft-margin formulation, which allows certain errors; and the Radial Basis Function (RBF) kernel, which builds nonlinear classifiers(20).

### Model performance

Each model was validated following a 5-fold cross-validation (CV) strategy given the size of the dataset. In the case of the SVM classifiers, a nested-CV was implemented consisting of a 5-fold inner CV to select the optimal parameters of the SVM. This inner loop follows a grid search for the regularization constant C ranging from 2-^4^ to 2^4^ both for soft-margin and RBF SVMs and a gamma γ ranging from 2^-4^ to 2^4^ for the RBF SVM. Once the optimal SVM model is selected during the inner CV, the outer CV is used to evaluate the classification performance. This nested CV offers an unbiased assessment of the model and prevents overestimation. Figure 1 illustrates the followed procedure. Furthermore, this whole procedure was repeated 100 times, to avoid bias and increase robustness.

For each model, we report the following measures of predictive performance: balanced accuracy (referred as accuracy (Acc) for the remainder of this work), sensitivity (Sens), specificity (Spec), positive (PPV) and negative predictive value (NPV), mean receiver operating characteristic (ROC) curves with 95% confidence intervals (CI) and area under the curve (AUC).

An important parameter of the GA is the length of each chromosome, that is, the length of the desired feature set. This parameter was set to 8 features. In practice, the selection of this parameter ought to be identified during the training step to avoid overfitting. Nonetheless, this step increases significantly the computational time of our proposed method. Regarding this computational time, a large initial population size to avoid premature convergence was prioritized, since it provides a better chance of significantly improving the feature selection process(21) and a high number of generations. Even so, the performance of our framework (Figure 1) for different combinations of this parameter was additionally explored to evaluate most selected features.

## Results

### Classification performance

Fifty four fatigued RA patients attended both visits. According to the fatigue scores at the second visit, 22 patients improved their fatigue levels and 32 remained with the same fatigue levels. Three different classifiers (LSLD, linear SVM and RBF SVM) were applied to each modality (sMRI, DTI and clinical) in order to distinguish between fatigue improvers and non-improvers. Figure 2, A) shows the ROC curves and corresponding AUC values for each classifier and modality during the CV step. All three classifiers performed similarly (< 3% difference), yet LSLD reached the highest accuracy in all cases. Classification performance using the sMRI features resulted in an accuracy of 64.3%, with 67.3% sensitivity and 61.2% specificity. DTI features reached 59.1% accuracy with 60% sensitivity and 58.3% specificity. The set of clinical features achieved 46.8% accuracy, 41.9% sensitivity and 51.8% specificity. **Table S4** provides detailed information of the classification performance for each algorithm and modality.

**Figure 2.**
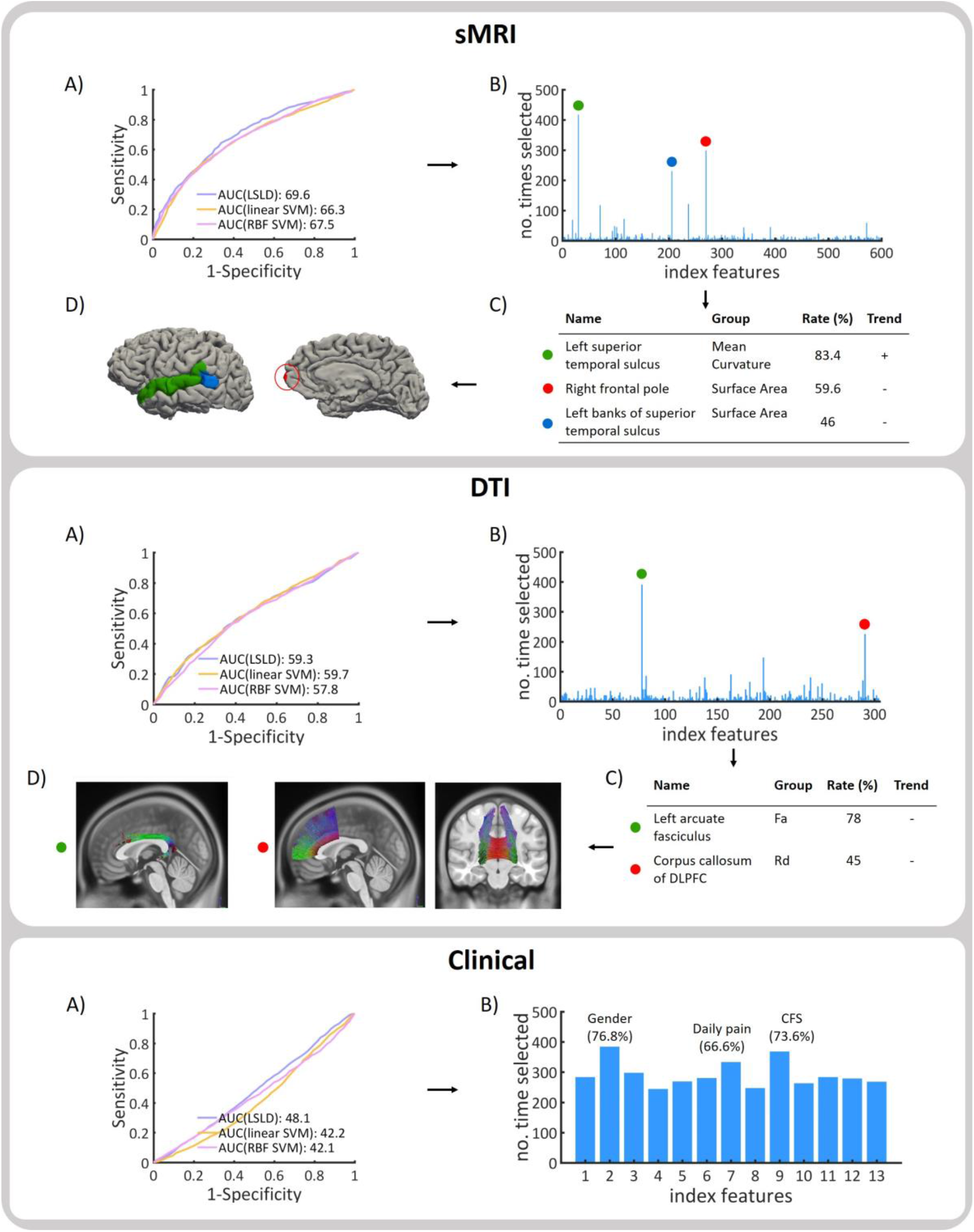
A) ROC curves and AUC values for three classifiers for 100 iterations of the CV procedure. B) Number of times each feature is selected during the feature selection for 100 iterations of the 5-fold CV procedure. C) Group: type of measure. Rate: frequency of repetition. Trend: +/- according to if the feature is increased/decreased in non-improvers compared to improvers of fatigue. Fa: fractional anisotropy. Rd: radial diffusivity. CFS: Chalder Fatigue Scale.

### Predictive features

The most selected features during the CV procedure across 100 iterations and thus, those features with the greatest predictive power between improvers and non-improvers of fatigue were extracted for each modality. Figure 2 displays these features, along with the percentage of selection during the CV and trend according to a 2 sample t-test analysis.

#### Structural MRI

Three features stood up for the 100 iterations of the CV process (Figure 2, sMRI, B-C), including the mean curvature of the left superior temporal sulcus (selected 83.4% of the times), the surface area of the right frontal pole (59.6%) and the surface area of the left banks of superior temporal sulcus (46%).

#### Diffusion Tensor Imaging

Two DTI features were consistently selected during the CV process (Figure 2, DTI, B-C). These are the fractional anisotropy of the left arcuate fasciculus (selected 78% of the times) and the radial diffusivity of the corpus callosum of the dorsal lateral prefrontal cortex (45%). Associated ROIs for each WM tract bundle according to the list of 76 WM tracts defined by Chen and colleagues(22) are listed in **Table S3**.

#### Clinical

Although no clinical feature clearly stood up in the CV process (Figure 2, Clinical, B), gender (76.8%), Chalder Fatigue Scale (73.6%) and daily pain (66.6%) measured during the first visit were the most selected ones.

### Varying the length of the “chromosome”

Figure 3, A) shows the classification accuracy at 95% CI during CV for three classifiers for each modality (sMRI, DTI and clinical) with the length of the chromosome during the feature selection step ranging from 1 to 20 features. Further measures of predictive performance and results for subgroups of features are summarized in supplementary material.

**Figure 3.**
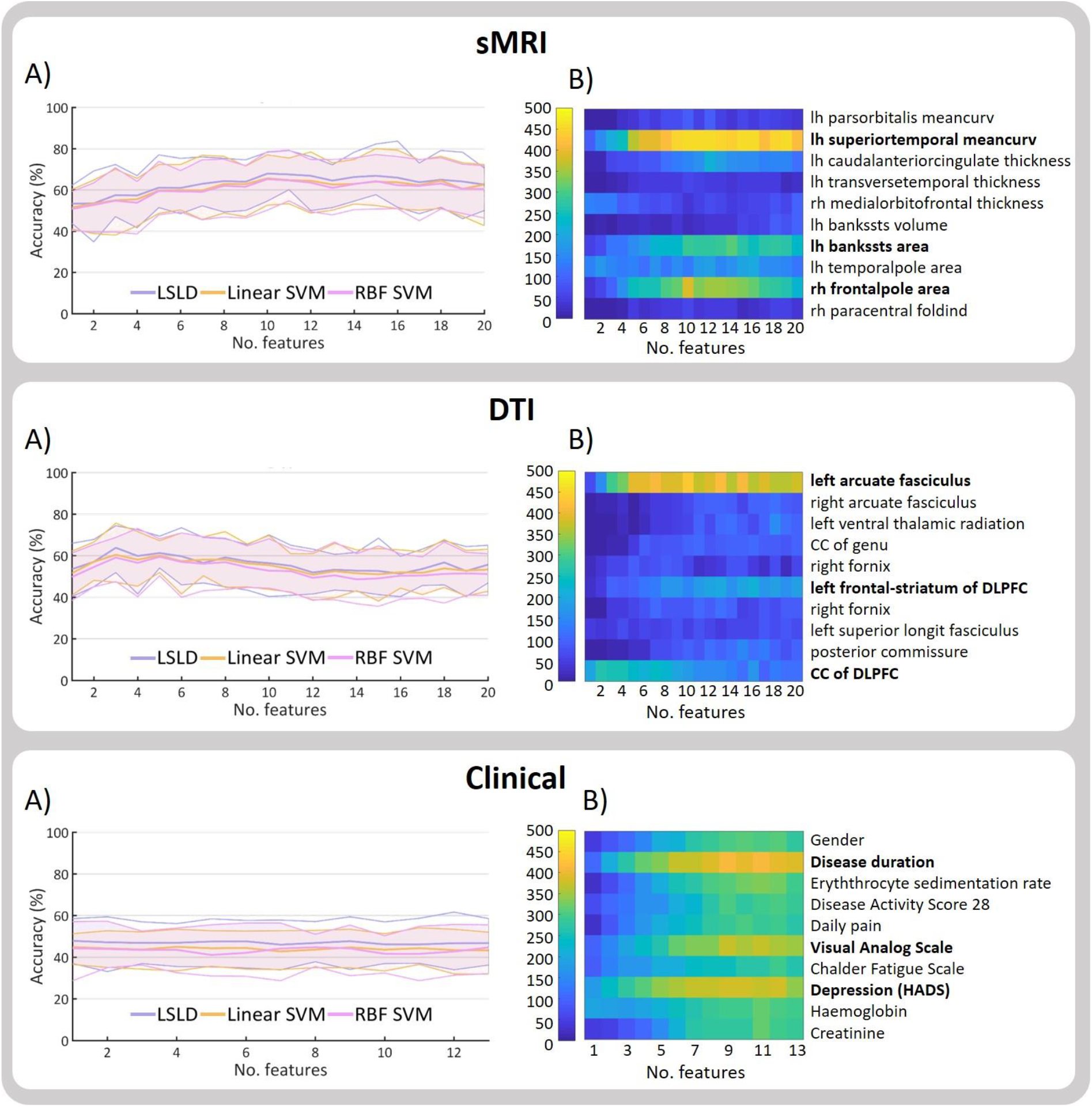
A) Accuracy with 95% CI for each classifier for different chromosome length (no. of features). B) Number of times (0-500) that a feature is selected during the 100 iterations of the 5-fold CV for different chromosome lengths for the LSLD (best performing classifier). For summarizing purposes, the ten most selected features for each modality are displayed. CC: corpus callosum. DLPFC: dorsolateral prefrontal cortex. HADS: Hospital Anxiety and Depression Scale. Lh: left hemisphere. Rh: right hemisphere.

Best performance for sMRI was achieved using 10 features as length of chromosome (Acc: 67.9%; Sens: 73.4%; Spec: 62.4%). The performance of the model decreased with chromosome lengths smaller than 8 features. Best performance for DTI modality was reached using 3 features (Acc: 63.8%; Sens: 66.5%; Spec: 61.1%). Highest accuracies for DTI were achieved with small chromosome lengths, ranging between 3 and 8 features. For clinical features, highest accuracy was reached using one feature (Acc: 47.9%; 47.4%; 48.3%). All results were in the chance level. LSLD was the classifier performing best for all modalities.

Number of times that each feature is selected during the 100 iterations of the 5-fold CV for each value of chromosome length was extracted (Figure 3, B). For the case of sMRI, the mean curvature of the left superior temporal sulcus was exceedingly the most selected in all cases, followed by the surface are of the right frontal pole and the surface area of the left banks of the superior temporal sulcus. For the DTI, the left arcuate fasciculus was clearly the most selected feature for all cases of chromosome length, followed by the corpus callosum of the dorsolateral prefrontal cortex (DLPFC) and the left frontal-striatum of the DLPFC. In the case of clinical features, the disease duration, visual analog scale and depression score as measured by the Hospital Anxiety and Depression Scale (HADS) were the most selected, although all features were selected at least once in different iterations.

## Discussion

The discovery of objective biomarkers of fatigue is critical to successful development of therapies. To date, current measures of fatigue are based on questionnaires which are after all, subjective measures. In addition, a prediction model of fatigue may support stratification of patients, saving time and frustration to both the clinician and patients, as being cost-effective for health services by a priori selecting therapeutic strategies which might be preferentially responsive for individual patients. Our results show the superiority of MR metrics in the prediction of fatigue changes, compared with commonly used clinical measures. This is the first study employing neuroimaging to predict fatigue outcome in rheumatoid arthritis subjects. In addition, it is the largest neuroimaging database in RA to date.

A machine learning approach was employed to predict improvement in fatigue in patients with RA and to identify those brain features with most discriminative power. Clinical, structural and diffusion brain features from MRI at baseline and 6 months apart, from 54 RA patients were assessed. Features based on ROI were used to reduce dimensionality and facilitate clinical interpretation. A classification procedure was performed to distinguish between improvers and non-improvers at baseline.

### Prediction of fatigue changes

All three classifiers performed very similarly in the discrimination of improvers and non-improvers of fatigue (<3% of difference), with LSLD performing best in all cases. Best prediction accuracy (64.3%) was reached using sMRI features, with 67.3% sensitivity and 61.2% specificity. Since sensitivity is greater than specificity, sMRI features seem to work better when predicting improvers than non-improvers of fatigue. With regard to DTI features, a modest accuracy of 59.1% was achieved, with 60% sensitivity and 58.3% specificity. Performance accuracy for the clinical set was in the chance level (46.8%), with 49.1% sensitivity and 51.8% specificity. Therefore, clinical features are not useful in this predictive problem, independently of the selection frequency during the feature selection step. Given that the clinician relies just on clinical data when prescribing a treatment for fatigue, and clinical data showed no predictive power, these results indicate a future asset in terms of therapeutic decision making support.

Whereas a chromosome length of 8 features was set during the feature selection step using the genetic algorithm, we also evaluated how variation of this parameter can modify the classification performance. For the sMRI set, accuracy increased from 64.3% to 67.9% when choosing 10 features. In the case of DTI features, accuracy raised from 59.1% to 63.8% when using 3 features instead of 8 features. Performance for the clinical set were all in the chance level.

### Identification of brain regions related to fatigue changes

#### Structural MRI

The following regions were highlighted in the discrimination between improvers and non-improvers of fatigue: curvature of the left superior temporal sulcus, the surface area of the right frontal pole and the surface area of the left bank superior temporal sulcus. It is worth noting that the frequency of these features was exceedingly high compared with the rest of the features. This may point out to a high relation between these brain regions and the fatigue pathology.

There is little work related to the investigation of the brain contributions to fatigue in RA patients using MRI. However, brain regions have been found to be related to fatigue in other chronic inflammatory diseases. Fatigue reduction was found to be correlated with cortical thinning of the superior temporal polysensory area in ankylosing spondylitis(23). In Chronic Fatigue Syndrome (CFS), a disorder which causes prominent fatigue, fatigue was significantly related with brain activity and superior temporal cortices (24). The frontal lobe has been associated with performance on a wide variety of tasks including multi-tasking and prospective memory. The only fatigue related RA study to date found greater levels of functional connectivity between the medial prefrontal gyri and Dorsal Attention Network (DAN) in those patients with higher levels of fatigue (25). Frontal lobe has also been reported in many studies of fatigue in other chronic diseases. Studies in Chronic Fatigue Syndrome reported correlations of fatigue with medial prefrontal regions (26), dorsolateral prefrontal and dorsomedial prefrontal cortex (26,27), lateral prefrontal cortex (27), right prefrontal cortex (28), right inferior frontal cortex(24) and superior frontal gyrus (29). Several studies in Multiple Sclerosis (MS) have linked fatigue with abnormalities in the frontal pole. Fatigued MS patients showed a significant atrophy of superior frontal gyrus compared with non-fatigued MS patients (31). Fatigue-related behavioural abnormalities in MS has also been associated with increased activation in frontal areas including superior, medial, middle and inferior regions (31). Furthermore, cognitive fatigue was related to right frontal lobe lesions in traumatic brain injury (33). The high number of reports related to this region suggests a strong link of the frontal lobe with cognitive fatigue in chronic inflammatory diseases.

#### Diffusion Tensor Imaging

Brain regions of greater structural integrity, as measured by increased FA, were identified within the fatigue improver group, concretely the left arcuate fasciculus. Reduced RD was found in corpus callosum of DLPFC within the non-improvers group.

Although few studies have employed DTI to study cognitive fatigue, there is still some evidence supporting our findings. Arcuate fasciculus has not been directly reported as related with fatigue in other studies. Nevertheless, it has been associated with symptoms which might have relation with fatigue, such as disease severity in CFS (34) or depression in MS (35). Dorsolateral prefrontal cortex (DLPFC) is related to executive functions such as working memory, cognitive flexibility, planning and reasoning. Decreased diffusivity in the DLPFC was found in traumatic brain injury (TBI) patients, another disease characterized by fatigue (36). Primary fatigued MS subjects showed regional atrophy in the DLPFC in an MRI study (37).

### Limitations and further research

Our predictive model has proven to identify fatigue changes and revealed those features consistently selected across different values of chromosome length. However, a limitation of our work is that we predefined this parameter. In future work, we will learn to identify the best value for the chromosome length as part of the training step. This is particularly important since as demonstrated, the classification performance vary according to the chromosome length.

An inherent limitation of this work lies in using the Chalder Fatigue Scale (CFS) to stratify subjects into improvers and non-improvers of fatigue, which is a self-rated score questionnaire. This tool gathers both physical and mental scores. Nonetheless, it may give a general overview rather than capturing the severity of fatigue in depth. Additionally, since CFS is a self-assessment questionnaire, the final fatigue score may be biased for the subjective individual perception of fatigue. Still, CFS has been the chosen tool in this work as it has been widely applied and validated in the current literature.

Whereas all patients were taking usual care (no interventions for fatigue), medication was different between them. Nonetheless, none of the patients were taking antidepressants, which are known to induce neurological changes. However, a slight variability might be included due to different types of medication. Other limitation is that although patients were stratified between improvers and non-improvers of fatigue, changes of fatigue were different between subjects. Whereas those patients reporting better scores in the CFS during the second visit were classified as fatigue improvers, some patients reported slight changes compared to others with greater changes. A regression strategy instead of a classification approach might provide greater clinically relevant information about the progression of each patient.

Given the complexity of collecting and scanning patients, imaging datasets are generally quite small. Several tens of scans cost thousands of pounds and require months to collect. Still, this is the biggest neuroimaging database of RA patients to date.

Our model of fatigue would strongly benefit from replication prior to use in different fatigue interventions. For example, it has been demonstrated that some patients experience a substantial improvement of fatigue following anti-TNF treatment(4). Other non-pharmacological fatigue interventions include Cognitive Behavioural Therapy (CBT) and physical exercise (38). This could be an approach to test our model or even to improve it.

During a fatigue intervention, greater changes of fatigue can be expected with the time, as opposed to the modest spontaneously fatigue changes of the current database. Since the methods followed in this work can be extended to other dataset, the resulting model after a fatigue intervention would be expected to be more robust and reliable.

## Data Availability

All data produced in the present work are contained in the manuscript.

## Acknowledgments

The authors thank Dr Mariella DAlesandro and Dr Maryam Alsyedalhashem for their assistance in recruiting the participants and preprocessing much of the data, the MRI-radiographers (B. Jagpal, K. Klaasen, N. Crouch and B. Maclennan), G. Buchan, T. Morris and D. Younie. Most important of all our thanks go to the participants for their time and effort. Research completed as part of MG’s PhD, funded by Sir Jules Thorn Charitable Trust with data collection funded by Pfizer.

