## Supplementary Material for "Brain predictors of fatigue in Rheumatoid Arthritis: a machine learning study"

**María Goñi, PhD**

Phone : +34 661237023

**List of features**

**Clinical**

Standard clinical characteristics included age, gender, weight and disease duration. Blood samples were taken and erythrocyte sedimentation rate (ESR), creatinine and haemoglobin measures extracted. In addition, disease activity was determined through the European League Against Rheumatism (EULAR) Disease Activity Score (DAS) (39). Other possible associations of fatigue were measured, such as daily pain (measured as 0 = no pain, 10 = severe pain), as well as anxiety and depression levels through the Hospital Anxiety and Depression Scale (HADS) (40). Fatigue was measured according to the Chalder Fatigue Scale (CFS) (11) and fatigue Visual Analogue Scale (VAS-F) (41).

Table S1. Clinical features for all subjects (RA), improvers and non-improvers of fatigue.

|  | **RA (n = 54)** | **Improvers**  **(n = 22)** | **Non-improvers**  **(n = 32)** |
| --- | --- | --- | --- |
| Age (years), mean ± std | 55 ± 11.4 | 54.9 ± 13.5 | 55 ± 10.1 |
| Male/Female | 13/41 | 7/15 | 6/26 |
| Disease duration (years), mean ± std | 11.5 ± 9.4 | 10.2 ± 6.6 | 12.5 ± 11 |
| Weight (Kg.), mean ± std | 74.2 ± 16.6 | 75.8 ± 15.9 | 73 ± 17.3 |
| DAS, mean ± std | 3.6 ± 1.3 | 3.5 ± 1.3 | 3.7 ± 1.3 |
| Daily pain, mean ± std | 3.8 ± 2.4 | 3.5 ± 2.1 | 4.1 ± 2.5 |
| Anxiety, mean ± std | 7.7 ± 4.2 | 8.7 ± 4.3 | 7 ± 4.1 |
| Depression, mean ± std | 6.9 ± 3.9 | 7.3 ± 3.7 | 6.6 ± 4.1 |
| VAS, mean ± std | 0.4 ± 0.2 | 0.4 ± 0.2 | 0.4 ± 0.2 |
| CFS, mean ± std | 9.2 ± 1.6 | 9.5 ± 1.4 | 8.9 ± 1.7 |
| ESR, (mm/hour) mean ± std | 19.2 ± 14.7 | 17.2 ± 15.5 | 20.6 ± 14.2 |
| Haemoglobin (g/L), mean ± std | 136.6 ± 11.5 | 138.1 ± 11.7 | 135.6 ± 11.4 |
| Creatinine (mg/dL), mean ± std | 65.3 ± 18.3 | 63.5 ± 12.9 | 66.5 ± 21.3 |
| DAS: Disease Activity Score. ESR: erythrocyte sedimentation rate. VAS: Visual Analogue Scale. CFS: Chalder Fatigue Scale | | | |

**Structural MRI ROIs**

Table S2. Cortical and subcortical parcellations extracted using FreeSurfer

| No. | ROI | Group | No. | ROI | Group |
| --- | --- | --- | --- | --- | --- |
| **1** | L_bankssts | Cortical | **61** | R_superiorfrontal | Cortical |
| **2** | L_caudalanteriorcingulate | Cortical | **62** | R_superiorparietal | Cortical |
| **3** | L_caudalmiddlefrontal | Cortical | **63** | R_superiortemporal | Cortical |
| **4** | L_cuneus | Cortical | **64** | R_supramarginal | Cortical |
| **5** | L_entorhinal | Cortical | **65** | R_frontalpole | Cortical |
| **6** | L_fusiform | Cortical | **66** | R_temporalpole | Cortical |
| **7** | L_inferiorparietal | Cortical | **67** | R_transversetemporal | Cortical |
| **8** | L_inferiortemporal | Cortical | **68** | R_insula | Cortical |
| **9** | L_isthmuscingulate | Cortical | **69** | Left-Lateral-Ventricle | Subcortical |
| **10** | L_lateraloccipital | Cortical | **70** | Left-Inf-Lat-Vent | Subcortical |
| **11** | L_lateralorbitofrontal | Cortical | **71** | Left-Cerebellum-White-Matter | Subcortical |
| **12** | L_lingual | Cortical | **72** | Left-Cerebellum-Cortex | Subcortical |
| **13** | L_medialorbitofrontal | Cortical | **73** | Left-Thalamus-Proper | Subcortical |
| **14** | L_middletemporal | Cortical | **74** | Left-Caudate | Subcortical |
| **15** | L_parahippocampal | Cortical | **75** | Left-Putamen | Subcortical |
| **16** | L_paracentral | Cortical | **76** | Left-Pallidum | Subcortical |
| **17** | L_parsopercularis | Cortical | **77** | 3rd-Ventricle | Subcortical |
| **18** | L_parsorbitalis | Cortical | **78** | 4th-Ventricle | Subcortical |
| **19** | L_parstriangularis | Cortical | **79** | Brain-Stem | Subcortical |
| **20** | L_pericalcarine | Cortical | **80** | Left-Hippocampus | Subcortical |
| **21** | L_postcentral | Cortical | **81** | Left-Amygdala | Subcortical |
| **22** | L_posteriorcingulate | Cortical | **82** | CSF | Subcortical |
| **23** | L_precentral | Cortical | **83** | Left-Accumbens-area | Subcortical |
| **24** | L_precuneus | Cortical | **84** | Left-VentralDC | Subcortical |
| **25** | L_rostralanteriorcingulate | Cortical | **85** | Left-vessel | Subcortical |
| **26** | L_rostralmiddlefrontal | Cortical | **86** | Left-choroid-plexus | Subcortical |
| **27** | L_superiorfrontal | Cortical | **87** | Right-Lateral-Ventricle | Subcortical |
| **28** | L_superiorparietal | Cortical | **88** | Right-Inf-Lat-Vent | Subcortical |
| **29** | L_superiortemporal | Cortical | **89** | Right-Cerebellum-White-Matter | Subcortical |
| **30** | L_supramarginal | Cortical | **90** | Right-Cerebellum-Cortex | Subcortical |
| **31** | L_frontalpole | Cortical | **91** | Right-Thalamus-Proper | Subcortical |
| **32** | L_temporalpole | Cortical | **92** | Right-Caudate | Subcortical |
| **33** | L_transversetemporal | Cortical | **93** | Right-Putamen | Subcortical |
| **34** | L_insula | Cortical | **94** | Right-Pallidum | Subcortical |
| **35** | R_bankssts | Cortical | **95** | Right-Hippocampus | Subcortical |
| **36** | R_caudalanteriorcingulate | Cortical | **96** | Right-Amygdala | Subcortical |
| **37** | R_caudalmiddlefrontal | Cortical | **97** | Right-Accumbens-area | Subcortical |
| **38** | R_cuneus | Cortical | **98** | Right-VentralDC | Subcortical |
| **39** | R_entorhinal | Cortical | **99** | Right-vessel | Subcortical |
| **40** | R_fusiform | Cortical | **100** | Right-choroid-plexus | Subcortical |
| **41** | R_inferiorparietal | Cortical | **101** | WM-hypointensities | Subcortical |
| **42** | R_inferiortemporal | Cortical | **102** | non-WM-hypointensities | Subcortical |
| **43** | R_isthmuscingulate | Cortical | **103** | Optic-Chiasm | Subcortical |
| **44** | R_lateraloccipital | Cortical | **104** | CC_Posterior | Subcortical |
| **45** | R_lateralorbitofrontal | Cortical | **105** | CC_Mid_Posterior | Subcortical |
| **46** | R_lingual | Cortical | **106** | CC_Central | Subcortical |
| **47** | R_medialorbitofrontal | Cortical | **107** | CC_Mid_Anterior | Subcortical |
| **48** | R_middletemporal | Cortical | **108** | CC_Anterior | Subcortical |
| **49** | R_parahippocampal | Cortical | **109** | BrainSegVolNotVent | Subcortical |
| **50** | R_paracentral | Cortical | **110** | BrainSegVol | Subcortical |
| **51** | R_parsopercularis | Cortical | **111** | lhCortexVol | Subcortical |
| **52** | R_parsorbitalis | Cortical | **112** | rhCortexVol | Subcortical |
| **53** | R_parstriangularis | Cortical | **113** | CortexVol | Subcortical |
| **54** | R_pericalcarine | Cortical | **114** | lhCorticalWhiteMatterVol | Subcortical |
| **55** | R_postcentral | Cortical | **115** | rhCorticalWhiteMatterVol | Subcortical |
| **56** | R_posteriorcingulate | Cortical | **116** | CorticalWhiteMatterVol | Subcortical |
| **57** | R_precentral | Cortical | **117** | SubCortGrayVol | Subcortical |
| **58** | R_precuneus | Cortical | **118** | TotalGrayVol | Subcortical |
| **59** | R_rostralanteriorcingulate | Cortical | **119** | SupraTentorialVol | Subcortical |
| **60** | R_rostralmiddlefrontal | Cortical | **120** | IntraCranialVol | Subcortical |

**DTI white matter tracts**

Table S3 gives details of the 76 major white matter tracts, categories and associated ROIs according to Chen et al (22).

Table S3. List of WM tract bundle with associated ROIs

| **No.** | **System** | **Subgroup** | **Name** | **Connected ROIs** | **Connected ROIs** |
| --- | --- | --- | --- | --- | --- |
| **1** | Association |  | L_AF | L_inferior frontal gyrus opercular part | L_superior temporal gyrus |
| **2** | Association |  | R_AF | R_inferior frontal gyrus opercular part | R_superior temporal gyrus |
| **3** | Association |  | L_cingulum of main body component | L_cingulate gyrus (anterior + middle parts) | L_cingulate gyrus posterior part |
| **4** | Association |  | R_cingulum of main body component | R_cingulate gyrus (anterior + middle parts) | R_cingulate gyrus posterior part |
| **5** | Association |  | L_cingulum of hippocampal component | L_cingulate gyrus posterior part | L_hippocampus |
| **6** | Association |  | R_cingulum of hippocampal component | R_cingulate gyrus posterior part | R_hippocampus |
| **7** | Association |  | L_frontal aslant tract | L_SMA | L_inferior frontal gyrus opercular part |
| **8** | Association |  | R_frontal aslant tract | R_SMA | R_inferior frontal gyrus opercular part |
| **9** | Association |  | L_fornix | L_mammillary body | L_hippocampus |
| **10** | Association |  | R_fornix | R_mammillary body | R_hippocampus |
| **11** | Association |  | L_IFOF | L_orbitofrontal gyrus | Occipital lobe |
| **12** | Association |  | R_IFOF | R_orbitofrontal gyrus | Occipital lobe |
| **13** | Association |  | L_ILF | L_temporal pole | Occipital lobe |
| **14** | Association |  | R_ILF | R_temporal pole | Occipital lobe |
| **15** | Association |  | L_perpendicular fasciculus | L_angular gyrus | L_temporal-parietal gyrus |
| **16** | Association |  | R_perpendicular fasciculus | R_angular gyrus | R_temporal-parietal gyrus |
| **17** | Association |  | L_SLF I | L_superior frontal gyrus | L_precuneus |
| **18** | Association |  | R_SLF I | R_superior frontal gyrus | R_precuneus |
| **19** | Association |  | L_SLF II | L_inferior frontal gyrus triangular part | L_middle occipital gyrus |
| **20** | Association |  | R_SLF II | R_inferior frontal gyrus triangular part | R_middle occipital gyrus |
| **21** | Association |  | L_SLF III | L_inferior frontal gyrus opercular part | L_angular gyrus |
| **22** | Association |  | R_SLF III | R_inferior frontal gyrus opercular part | R_angular gyrus |
| **23** | Association |  | L_stria terminalis | L_septal nuclei | L_amygdala |
| **24** | Association |  | R_stria terminalis | R_septal nuclei | R_amygdala |
| **25** | Association |  | L_UF | L_orbitofrontal gyrus | L_superior temporal pole |
| **26** | Association |  | R_UF | R_orbitofrontal gyrus | R_superior temporal pole |
| **27** | Projection | CST | L_CST of hand | Brain stem | L_primary motor cortex of hand component |
| **28** | Projection | CST | R_CST of hand | Brain stem | R_primary motor cortex of hand component |
| **29** | Projection | CST | L_CST of trunk | Brain stem | L_primary motor cortex of trunk component |
| **30** | Projection | CST | R_CST of trunk | Brain stem | R_primary motor cortex of trunk component |
| **31** | Projection | CST | L_CST of mouth | Brain stem | L_primary motor cortex of mouth component |
| **32** | Projection | CST | R_CST of mouth | Brain stem | R_primary motor cortex of mouth component |
| **33** | Projection | CST | L_CST of toe | Brain stem | L_primary motor cortex of toe component |
| **34** | Projection | CST | R_CST of toe | Brain stem | R_primary motor cortex of toe component |
| **35** | Projection | CST | L_CST of geniculate fibers | Brain stem | L_primary motor cortex of throat component |
| **36** | Projection | CST | R_CST of geniculate fibers | Brain stem | R_primary motor cortex of throat component |
| **37** | Projection | FS | L_FS of OFC | L_striatum (putamen + caudate) | L_orbitofrontal gyrus |
| **38** | Projection | FS | R_FS of OFC | R_striatum (putamen + caudate) | R_orbitofrontal gyrus |
| **39** | Projection | FS | L_FS of VLPFC | L_striatum (putamen + caudate) | L_inferior frontal gyrus + middle frontal gyrus |
| **40** | Projection | FS | R_FS of VLPFC | R_striatum (putamen + caudate) | R_inferior frontal gyrus + middle frontal gyrus |
| **41** | Projection | FS | L_FS of DLPFC | L_striatum (putamen + caudate) | L_medial frontal gyrus + superior frontal gyrus |
| **42** | Projection | FS | R_FS of DLPFC | R_striatum (putamen + caudate) | R_medial frontal gyrus + superior frontal gyrus |
| **43** | Projection | FS | L_FS of precentral gyrus | L_striatum (putamen + caudate) | L_precentral gyrus |
| **44** | Projection | FS | R_FS of precentral gyrus | R_striatum (putamen + caudate) | R_precentral gyrus |
| **45** | Projection | L_Medial lemniscus | Brain stem | L_thalamus | 45 |
| **46** | Projection | R_Medial lemniscus | Brain stem | R_thalamus | 46 |
| **47** | Projection | TR | L_TR of VLPFC | L_thalamus | L_orbitofrontal gyrus + middle frontal gyrus + inferior frontal gyrus |
| **48** | Projection | TR | R_TR of VLPFC | R_thalamus | R orbitofrontal gyrus + middle frontal gyrus + inferior frontal gyrus |
| **49** | Projection | TR | L_TR of DLPFC | L_thalamus | L_medial frontal gyrus + superior frontal gyrus + SMA |
| **51** | Projection | TR | L_TR of precentral gyrus | L_thalamus | L_precentral gyrus |
| **52** | Projection | TR | R_TR of precentral gyrus | R_thalamus | R_precentral gyrus |
| **53** | Projection | TR | L_TR of postcentral gyrus | L_thalamus | L_postcentral gyrus |
| **54** | Projection | TR | R_TR of postcentral gyrus | R_thalamus | R_postcentral gyrus |
| **55** | Projection | TR | L_TR of auditory nerve | L_thalamus | L_Heschl’s gyrus |
| **56** | Projection | TR | R_TR of auditory nerve | R_thalamus | R_Heschl’s gyrus |
| **57** | Projection | TR | L_TR of optic radiation | L_thalamus | L_superior occipital gyrus |
| **58** | Projection | TR | R_TR of optic radiation | R_thalamus | R_superior occipital gyrus |
| **59** | Commisure | Anterior commissure | Front column of fornix | Bilateral cerebral hemispheres | 59 |
| **60** | Commisure | Posterior commisure | Dorsal aspect of the upper end of the cerebral aqueduct | Bilateral cerebral hemispheres | 60 |
| **61** | Commisure | CC | CC of genu | L_orbitofrontal gyrus | R_orbitofrontal gyrus |
| **62** | Commisure | CC | CC of DLPFC | L_medial frontal gyrus + superior frontal gyrus | R_medial frontal gyrus + superior frontal gyrus |
| **63** | Commisure | CC | CC of VLPFC | L_inferior frontal gyrus + middle frontal gyrus | R_inferior frontal gyrus + middle frontal gyrus |
| **64** | Commisure | CC | CC of SMA | L_supplementary motor areas | R_supplementary motor areas |
| **65** | Commisure | CC | CC of precentral gyrus | L_precentral gyrus | R_precentral gyrus |
| **66** | Commisure | CC | CC of paracentral lobule | L_paracentral lobules | R_paracentral lobules |
| **67** | Commisure | CC | CC of inferior parietal lobule | L_inferior parietal lobules | R_inferior parietal lobules |
| **68** | Commisure | CC | CC of postcentral gyrus | L_postcentral gyrus | R_postcentral gyrus |
| **69** | Commisure | CC | CC of superior parietal lobule | L_superior parietal lobules | R_superior parietal lobules |
| **70** | Commisure | CC | CC of superior temporal gyrus | L_superior temporal gyrus | R_superior temporal gyrus |
| **71** | Commisure | CC | CC of middle temporal gyrus | L_middle temporal gyrus | R_middle temporal gyrus |
| **72** | Commisure | CC | CC of temporal pole | L_temporal poles | R_temporal poles |
| **73** | Commisure | CC | CC of hippocampus | L_hippocampus | R_hippocampus |
| **74** | Commisure | CC | CC of amygdale | L_amygdala | R_amygdala |
| **75** | Commisure | CC | CC of precuneus | L_precuneus | R_precuneus |
| **76** | Commisure | CC | CC of splenium | L_occipital lobe | R_occipital lobe |
| AF:arcuate fasciculus; CC: corpus callosum; CST: corticospinal tract; DLPFC: dorsal lateral prefrontal cortex; FS: frontal-striatum; IFOF: inferior frontal occipital fasciculus; ILF: inferior longitudinal fasciculus; L: left; OFC: orbitofrontal cortex; R: right; SLF: superior longitudinal fasciculus; SMA: supplementary motor area; TR: thalamic radiation; UF: uncinate fasciculus; VLPFC: ventral lateral prefrontal cortex. | | | | | |

**Results**

**Main results**

The following table summarize the results for each modality when selecting a chromosome length of 8 features during the feature selection step.

Table S4. Cross-validation classification performances for each modality (sMRI, DTI and clinical features) for three different classifiers

|  |  |  |  | **% (95% CI)** | **LSLD** | **Linear SVM** | **RBF SVM** |
| --- | --- | --- | --- | --- | --- | --- | --- |
| **sMRI** | | | | **Acc** | 64.3 (49.3; 75.3) | 62.9 (48.7; 76.3) | 62 (46.9; 74.9) |
|  |  |  |  | **Sens** | 67.3 (45.5; 81.8) | 55.6 (31.8; 72.7) | 51.5 (27.3; 68.2) |
|  |  |  |  | **Spec** | 61.2 (40.6; 75) | 70.2 (46.9; 84.4) | 72.5 (43.8; 90.6) |
|  |  |  |  | **PPV** | 54.6 (40; 66.7) | 56.6 (38.9; 75) | 56.9 (37.9; 78.6) |
|  |  |  |  | **NPV** | 73.3 (58.6; 84.6) | 69.7 (58.3; 81.3) | 68.5 (56; 78.1) |
|  |  |  |  | **AUC** | 69.6 (59.1; 86.2) | 66.3 (51.6; 80.5) | 67.5 (53.3; 84) |
| **DTI** | | | | **Acc** | 59.1 (45; 68.1) | 58.1 (44.7; 71.6) | 56.8 (43.8; 68.4) |
|  |  |  |  | **Sens** | 60 (40; 80) | 46.5 (20; 65) | 44.2 (25; 65) |
|  |  |  |  | **Spec** | 58.3 (46.9; 75) | 69.7 (56.3; 90.6) | 69.5 (59.4; 81.3) |
|  |  |  |  | **PPV** | 47.3 (33.3; 57.9) | 49.3 (31.6; 76.9) | 47.3 (29.4; 62.5) |
|  |  |  |  | **NPV** | 70.3 (57.1; 81.8) | 67.7 (57.6; 77.4) | 66.8 (57.1; 76.7) |
|  |  |  |  | **AUC** | 59.3 (45.5; 75.2) | 59.7 (44.5; 74.8) | 57.8 (43.1; 75.6) |
| **Clinical** | | | | **Acc** | 46.8 (37.8; 57.1) | 43.6 (34.9; 52.8) | 44.8 (35.5; 51) |
|  |  |  |  | **Sens** | 41.9 (27.3; 59.1) | 18.3 (4.6; 31.8) | 34.1 (27.3; 45.5) |
|  |  |  |  | **Spec** | 51.8 (31.3; 68.8) | 69 (46.9; 87.5) | 55.5 (43.8; 65.6) |
|  |  |  |  | **PPV** | 37.4 (28; 50) | 28.3 (9.1; 50) | 34.5 (25; 42.1) |
|  |  |  |  | **NPV** | 56.3 (48.2; 65.6) | 55 (48.3; 60.9) | 55 (46.7; 60) |
|  |  |  |  | **AUC** | 48.1 (36.8; 62.2) | 42.2 (32; 55) | 42.1 (35.4; 46.5) |

**Varying the length of the chromosome**

*Structural MRI*

Figure S1 shows the classification accuracy at 95% CI during the CV for the three classifiers for each subset of sMRI features. For visualization purposes the case of maximum accuracy is highlighted with a square. For each case, further measures of predictive performance are summarized in Table S5.

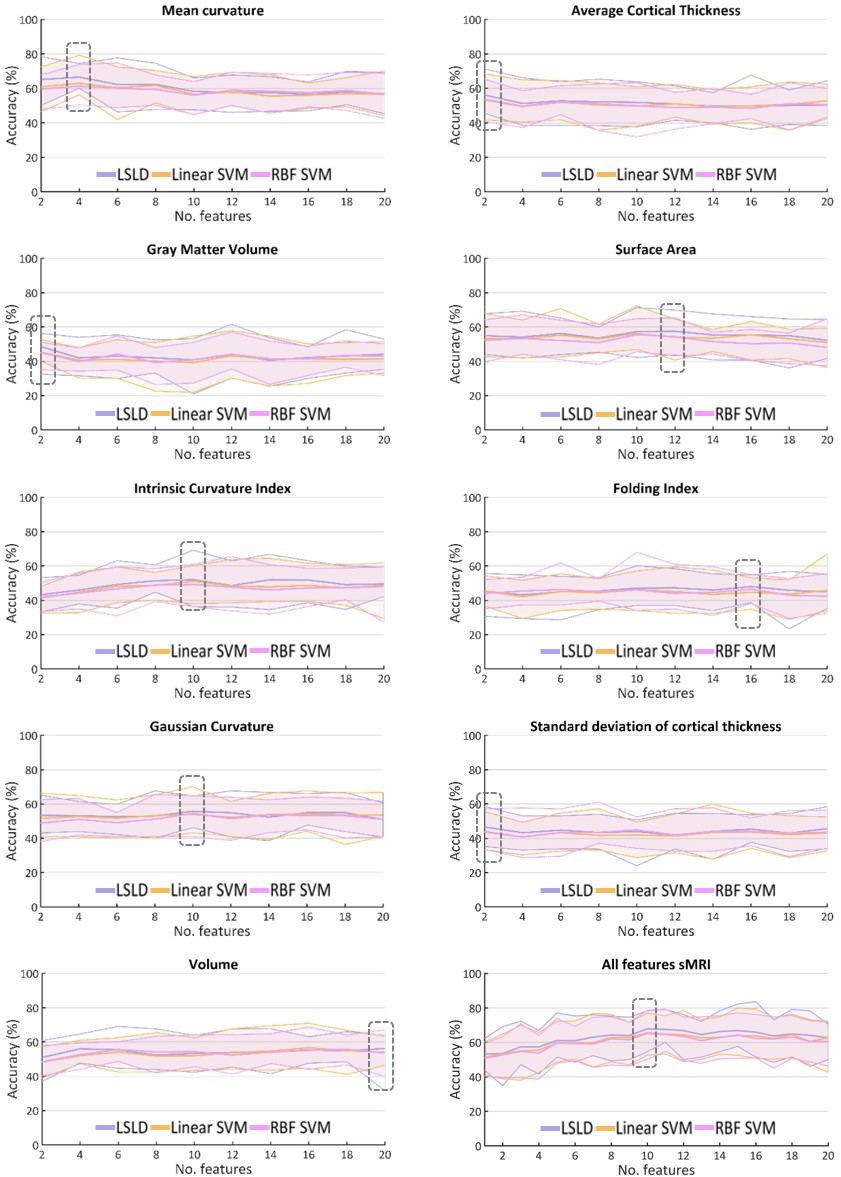

Figure S1. Accuracy with 95% CI for each classifier for different chromosome length (no. of features) for each subset of sMRI features. Best classification performance is marked in a square.

Table S5. Classification performance for the case of maximum accuracy when varying the chromosome length (No. feat) for each subset of sMRI features.

|  |  |  |  | **% (95% CI)** | **LSLD** | **Linear SVM** | **RBF SVM** |
| --- | --- | --- | --- | --- | --- | --- | --- |
| **Mean**  **curvature** | | | | **No. feat.** | 4 | 4 | 4 |
|  |  |  |  | **Acc** | 66.5 (60.1; 74.4) | 62.9 (56.3; 79.3) | 61.3 (50.3; 73.9) |
|  |  |  |  | **Sens** | 70.7 (54.5; 86.4) | 54.8 (31.8; 77.3) | 50 (31.8; 72.7) |
|  |  |  |  | **Spec** | 62.3 (53.1; 68.8) | 71.1 (59.4; 81.3) | 72.5 (62.5; 81.3) |
|  |  |  |  | **PPV** | 56.3 (51.9; 63) | 56.5 (47.8; 73.9) | 55.5 (41.2; 70) |
|  |  |  |  | **NPV** | 75.8 (67.7; 87) | 69.8 (63.4; 83.9) | 68 (59.5; 80) |
|  |  |  |  | **AUC** | 70.2 (63.9; 81.4) | 67.2 (60.4; 80.1) | 67.3 (57.2; 81.3) |
| **Average**  **cortical**  **thickness** | | | | **No. feat.** | 2 | 2 | 2 |
|  |  |  |  | **Acc** | 56.2 (45.5; 71.3) | 53.3 (41.8; 68.5) | 53.6 (40.9; 65.3) |
|  |  |  |  | **Sens** | 62.1 (40.9; 86.4) | 44.8 (27.3; 72.7) | 42.3 (22.7; 68.2) |
|  |  |  |  | **Spec** | 50.3 (37.5; 68.8) | 61.7 (46.9; 78.1) | 65 (50; 78.1) |
|  |  |  |  | **PPV** | 46.3 (36; 58.3) | 44.4 (30; 60) | 45.4 (30.4; 58.8) |
|  |  |  |  | **NPV** | 66 (54.6; 85.7) | 62.2 (52.9; 75.9) | 62.2 (51.6; 74.1) |
|  |  |  |  | **AUC** | 56.3 (44.7; 72) | 54.5 (43.3; 66.1) | 54.3 (41.1; 65.3) |
| **Gray matter volume** | | | | **No. feat.** | 2 | 2 | 2 |
|  |  |  |  | **Acc** | 48.6 (32.4; 56.3) | 45.2 (40.2; 52.7) | 45.1 (35.1; 51) |
|  |  |  |  | **Sens** | 48.9 (27.3; 59.1) | 25.2 (9.1; 40.9) | 24.8 (4.6; 36.4) |
|  |  |  |  | **Spec** | 48.3 (37.5; 62.5) | 65.2 (50; 78.1) | 65.5 (53.1; 78.1) |
|  |  |  |  | **PPV** | 39.4 (23.1; 47.8) | 32.9 (22.2; 46.2) | 32.2 (8.3; 42.1) |
|  |  |  |  | **NPV** | 57.8 (42.9; 64.5) | 55.8 (51.5; 61) | 55.9 (50; 60) |
|  |  |  |  | **AUC** | 45.1 (29.3; 52.4) | 43.5 (31.7; 51.6) | 46.3 (39.4; 55.4) |
| **Surface area** | | | | **No. feat.** | 12 | 10 | 10 |
|  |  |  |  | **Acc** | 57.5 (43.8; 69.9) | 56.2 (46.9; 72.3) | 55.5 (45.6; 64.8) |
|  |  |  |  | **Sens** | 62.5 (40.9; 81.8) | 49.6 (36.4; 72.7) | 44.8 (27.3; 63.6) |
|  |  |  |  | **Spec** | 52.5 (37.5; 68.8) | 62.8 (43.8; 78.1) | 66.3 (43.8; 78.1) |
|  |  |  |  | **PPV** | 47.7 (35.5; 58.6) | 48 (37.9; 64) | 48 (35; 60) |
|  |  |  |  | **NPV** | 67.2 (52.2; 80) | 64.5 (56; 79.3) | 63.7 (55.9; 70.6) |
|  |  |  |  | **AUC** | 59.5 (41.8; 75.1) | 59.4 (47.7; 76.9) | 59.8 (51; 69.6) |
| **Intrinsic curvature index** | | | | **No. feat.** | 10 | 10 | 10 |
|  |  |  |  | **Acc** | 52 (36.2; 69.2) | 51.1 (37.9; 60.1) | 49.2 (36.2; 60.9) |
|  |  |  |  | **Sens** | 53.2 (31.8; 72.7) | 42.1 (22.7; 59.1) | 36.8 (18.2; 54.6) |
|  |  |  |  | **Spec** | 50.8 (40.6; 65.6) | 60.2 (43.8; 68.8) | 61.6 (40.6; 78.1) |
|  |  |  |  | **PPV** | 42.4 (26.9; 59.3) | 41.7 (25; 52.2) | 39.6 (23.5; 55) |
|  |  |  |  | **NPV** | 61.5 (46.4; 77.8) | 60.3 (50; 67.7) | 58.6 (46.4; 67.7) |
|  |  |  |  | **AUC** | 56.9 (46.7; 69.7) | 54.3 (42.3; 66.2) | 48.8 (38.6; 64.1) |
| **Folding Index** | | | | **No. feat.** | 16 | 10 | 10 |
|  |  |  |  | **Acc** | 48 (38.6; 54.7) | 46.2 (34; 57.1) | 49.2 (36.2; 60.9) |
|  |  |  |  | **Sens** | 47.1 (27.3; 59.1) | 40 (27.3; 50) | 36.8 (18.2; 54.6) |
|  |  |  |  | **Spec** | 48.9 (37.5; 59.4) | 52.3 (34.4; 68.8) | 61.6 (40.6; 78.1) |
|  |  |  |  | **PPV** | 38.8 (27.3; 45.8) | 36.9 (24; 50) | 35.2 (20; 50) |
|  |  |  |  | **NPV** | 57.4 (48; 63.3) | 55.6 (44.8; 64.7) | 56.7 (50; 62.5) |
|  |  |  |  | **AUC** | 48.8 (39.5; 60) | 44.7 (29.7; 60.5) | 48.8 (38.6; 64.1) |
| **Gaussian Curvature** | | | | **No. feat.** | 10 | 16 | 10 |
|  |  |  |  | **Acc** | 55.7 (46.2; 64.6) | 54.2 (44; 67.8) | 54.5 (40.9; 64.8) |
|  |  |  |  | **Sens** | 57.5 (36.4; 72.7) | 50.7 (31.8; 63.6) | 41.8 (31.8; 54.6) |
|  |  |  |  | **Spec** | 53.9 (40.6; 65.6) | 57.7 (43.8; 71.9) | 67.2 (50; 78.1) |
|  |  |  |  | **PPV** | 46 (37; 56) | 45.4 (33.3; 60.9) | 47.2 (30.4; 61.1) |
|  |  |  |  | **NPV** | 65.2 (55.6; 73.9) | 62.9 (54.6; 74.2) | 62.6 (51.6; 70.6) |
|  |  |  |  | **AUC** | 57.2 (35.2; 72.9) | 54.8 (44.3; 68) | 54.6 (36.7; 68) |
| **Standard deviation of cortical thickness** | | | | **No. feat.** | 2 | 16 | 10 |
|  |  |  |  | **Acc** | 46.5 (35.2; 58.5) | 43.7 (34.1; 54.6) | 44.9 (34.1; 52.4) |
|  |  |  |  | **Sens** | 46.6 (18.2; 63.6) | 37.1 (13.6; 59.1) | 34.1 (18.2; 50) |
|  |  |  |  | **Spec** | 46.6 (25; 62.5) | 50.3 (37.5; 59.4) | 55.6 (43.8; 68.8) |
|  |  |  |  | **PPV** | 37.2 (22.2; 50) | 33 (18.8; 44.8) | 34.3 (20; 43.5) |
|  |  |  |  | **NPV** | 55.9 (40; 66.7) | 54.1 (46.7; 64) | 55.1 (47.1; 61.3) |
|  |  |  |  | **AUC** | 42.7 (28.3; 52.1) | 42.3 (34.4; 54.3) | 40.5 (23.4; 50.7) |
| **Volume** | | | | **No. feat.** | 20 | 16 | 18 |
|  |  |  |  | **Acc** | 56.2 (31.5; 63.8) | 56.6 (44.6; 70.9) | 55.7 (46.6; 64.1) |
|  |  |  |  | **Sens** | 57.7 (31.8; 72.7) | 48 (36.4; 63.6) | 36.1 (18.2; 54.6) |
|  |  |  |  | **Spec** | 54.7 (31.3; 62.5) | 65.3 (43.8; 78.1) | 75.3 (62.5; 87.5) |
|  |  |  |  | **PPV** | 46.7 (24.1; 53.6) | 49.4 (35.7; 66.7) | 50.7 (33.3; 66.7) |
|  |  |  |  | **NPV** | 65.4 (40; 73.1) | 64.4 (53.9; 75.8) | 63.2 (57.1; 69.4) |
|  |  |  |  | **AUC** | 56.4 (42.5; 65.8) | 57.5 (40.6; 66.1) | 55 (44.6; 61.5) |
| **All features sMRI** | | | | **No. feat.** | 10 | 10 | 10 |
|  |  |  |  | **Acc** | 67.9 (54.6; 78.3) | 65.6 (52.6; 77) | 65.2 (50.1; 78.6) |
|  |  |  |  | **Sens** | 73.4 (59.1; 90.9) | 60.1 (40.9; 77.3) | 58.3 (36.4; 77.3) |
|  |  |  |  | **Spec** | 62.4 (50; 81.3) | 71 (59.4; 84.4) | 72.1 (59.4; 87.5) |
|  |  |  |  | **PPV** | 57.7 (44.8; 71.4) | 59.1 (44.4; 72.7) | 59.2 (40.9; 77.8) |
|  |  |  |  | **NPV** | 77.5 (64; 91.3) | 72.3 (61.3; 82.8) | 71.7 (59.4; 81.8) |
|  |  |  |  | **AUC** | 73.2 (59.9; 87.4) | 71.7 (53.8; 87.8) | 71.6 (55.5; 88.8) |

*Diffusion Tensor Imaging*

Figure S2 shows the classification accuracy at 95% CI during the CV for the three classifiers for each subset of sMRI features. For visualization purposes the case of maximum accuracy is highlighted with a square. For each case, further measures of predictive performance are summarized in Table S6.

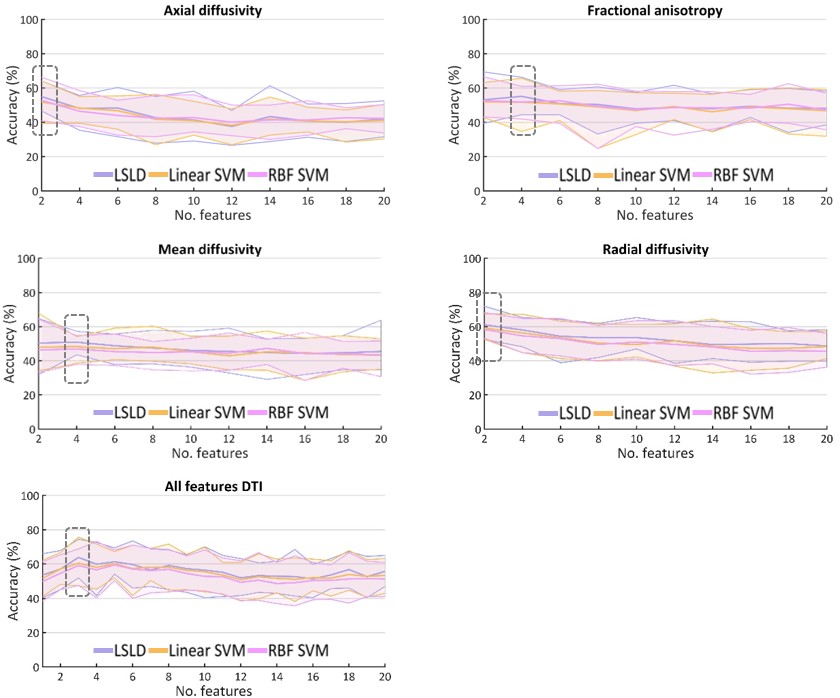

Figure S2. Accuracy with 95% CI for each classifier for different chromosome length (no. of features) for each subset of DTI features. Best classification performance is marked in a square.

Table S6. Classification performance for the case of maximum accuracy when varying the chromosome length (No. feat) for each subset of DTI features.

|  |  |  |  | **% (95% CI)** | **LSLD** | **Linear SVM** | **RBF SVM** |
| --- | --- | --- | --- | --- | --- | --- | --- |
| **Axial diffusivity** | | | | **No. feat.** | 2 | 2 | 2 |
|  |  |  |  | **Acc** | 54.9 (46.6; 64.1) | 51.8 (39.4; 64.1) | 52.8 (40.9; 66.3) |
|  |  |  |  | **Sens** | 58 (40; 75) | 30 (10; 50) | 27.8 (10; 55) |
|  |  |  |  | **Spec** | 51.9 (40.6; 65.6) | 73.6 (62.5; 87.5) | 77.8 (59.4; 96.9) |
|  |  |  |  | **PPV** | 42.9 (34.8; 50) | 40.8 (16.7; 60) | 44.9 (18.2; 69.2) |
|  |  |  |  | **NPV** | 66.6 (58.6; 77.3) | 62.9 (55; 71.4) | 63.4 (56.1; 71.8) |
|  |  |  |  | **AUC** | 56.6 (41.7; 69.8) | 54.2 (43; 67.2) | 51.2 (39.5; 65) |
| **Fractional anisotropy** | | | | **No. feat.** | 4 | 2 | 2 |
|  |  |  |  | **Acc** | 55.1 (44.4; 66.3) | 51.9 (42.8; 63.1) | 52.6 (43.1; 66.6) |
|  |  |  |  | **Sens** | 54.8 (30; 70) | 33 (20; 50) | 38 (25; 55) |
|  |  |  |  | **Spec** | 55.5 (43.8; 68.8) | 70.8 (56.3; 84.4) | 67.2 (56.3; 78.1) |
|  |  |  |  | **PPV** | 43.4 (31.6; 53.8) | 41.8 (26.7; 60) | 42.1 (30; 61.1) |
|  |  |  |  | **NPV** | 66.4 (56; 76.9) | 62.8 (56.8; 70.3) | 63.4 (56.3; 73.5) |
|  |  |  |  | **AUC** | 52.6 (37.2; 66.4) | 51.8 (37; 68.3) | 52.6 (37.5; 71.6) |
| **Mean diffusivity** | | | | **No. feat.** | 4 | 4 | 14 |
|  |  |  |  | **Acc** | 50.8 (43.4; 57.2) | 48.3 (38.4; 54.1) | 47.2 (37.8; 52.5) |
|  |  |  |  | **Sens** | 50.8 (40; 65) | 29 (10; 40) | 20.5 (5; 40) |
|  |  |  |  | **Spec** | 50.9 (40.6; 62.5) | 67.5 (46.9; 81.3) | 73.9 (56.3; 87.5) |
|  |  |  |  | **PPV** | 39.3 (32; 45.8) | 36 (20; 46.2) | 32.4 (15.4; 44.4) |
|  |  |  |  | **NPV** | 62.4 (55.6; 68) | 60.2 (51.7; 64.1) | 59.8 (53.9; 63.2) |
|  |  |  |  | **AUC** | 50.5 (38.9; 59.1) | 49.2 (35.8; 57.2) | 41.9 (24.2; 50.9) |
| **Radial diffusivity** | | | | **No. feat.** | 2 | 2 | 2 |
|  |  |  |  | **Acc** | 61 (52.5; 71.9) | 59.1 (52.5; 67.5) | 58.3 (53.1; 68.1) |
|  |  |  |  | **Sens** | 62 (45; 75) | 37.8 (20; 60) | 34.5 (20; 55) |
|  |  |  |  | **Spec** | 60 (50; 71.9) | 80.5 (65.6; 90.6) | 82 (68.8; 90.6) |
|  |  |  |  | **PPV** | 49.4 (40.7; 60) | 55.2 (42.1; 66.7) | 54.8 (44.4; 70) |
|  |  |  |  | **NPV** | 71.6 (64; 81.5) | 67.5 (63.2; 75) | 66.8 (63.4; 74.3) |
|  |  |  |  | **AUC** | 61.1 (49.5; 70.5) | 59.7 (46.7; 66.9) | 58.7 (45.3; 68.8) |
| **All features DTI** | | | | **No. feat.** | 3 | 3 | 5 |
|  |  |  |  | **Acc** | 63.8 (51.9; 74.4) | 60.4 (47.2; 75.6) | 59.4 (50.3; 68.1) |
|  |  |  |  | **Sens** | 66.5 (55; 80) | 48.5 (35; 70) | 47 (35; 60) |
|  |  |  |  | **Spec** | 61.10 (43.8; 75) | 72.4 (56.3; 84.4) | 72 (59.4; 90.6) |
|  |  |  |  | **PPV** | 52 (40; 63.6) | 52.6 (35; 70) | 51.8 (38.9; 72.7) |
|  |  |  |  | **NPV** | 74.4 (63.6; 84.6) | 69.3 (59.4; 81.3) | 68.4 (61.8; 75) |
|  |  |  |  | **AUC** | 63.1 (47.7; 74.7) | 61.8 (42.8; 74.8) | 60.8 (49.5; 71.6) |
